# Substance use and treatment utilization patterns of working-age American men who were not in employment, education, or training (NEET) during the COVID-19 pandemic

**DOI:** 10.64898/2025.12.18.25342578

**Authors:** Wayne Kepner, Keith Humphreys

**Author notes:** Corresponding Author: Wayne Kepner, PhD, MPH Department of Psychiatry and Behavioral Sciences, Stanford University School of Medicine, 1070 Arastradero Rd Stanford, Palo Alto, CA, United States, 94305.

## Abstract

**Background:** A growing population of working-aged men are not in employment, education, or training (NEET). The COVID-19 pandemic increased rates of substance use disorders (SUDs) and affected treatment seeking in the general population, but the COVID era substance use patterns among NEET men are unknown.

**Methods:** We estimated the prevalence and correlates of NEET status among working-aged (18–64) men using data from the 2022 National Survey on Drug Use and Health, a nationally representative survey of non-institutionalized individuals in the United States. We developed logistic regression models to examine associations between NEET status and substance use behaviors and treatment engagement, adjusted for sociodemographic factors.

**Results:** An estimated 11.1% of working-aged men were NEET in 2022 representing 10.6 million individuals. NEET men were significantly more likely to be older, have lower income, be unmarried, and have lower educational attainment and be Non-Hispanic Black compared to non-NEET men. After adjusting for sociodemographic factors, NEET status was significantly associated with higher odds of prescription tranquilizer/sedative use disorder (aOR = 3.54, 1.97-6.37), methamphetamine use disorder (aOR = 3.10, 95% CI: 1.82-5.28), and prescription pain reliever use disorder (aOR = 2.88, 1.82-4.53), while being inversely associated with alcohol use disorder (aOR = 0.68, 0.54-0.85).

**Conclusion:** More than 1 in 10 working-aged men were NEET in 2022. Adjusted models showed higher odds of past-year SUDs but lower rates of alcohol use disorder. Targeted interventions should include age-appropriate, culturally tailored, and substance-specific treatment programs to improve public health.

## 1. Introduction

The decline in American male workforce participation has created a growing population of men who are ‘not in employment, education or training (i.e., ‘NEET’); estimates suggest that 11% of prime-aged men (aged 25-54) were not participating in the labor force in 2019 (1, 2).

Traditional metrics of unemployment typically capture active jobseekers; it excludes individuals who are long-term unemployed, not looking for work, and at risk of permanent workforce exclusion. Thus, the NEET concept is the central focus of this study because it provides a broader framework that includes such individuals.

The COVID-19 pandemic created disruptions to both employment and substance use patterns among American men. During the pandemic, labor force participations rates among prime-aged men (25–54) dropped to 86.4% (in April of 2020), meaning that 13.6% were not in the labor force (1). COVID-19 and measures associated with it (e.g., social distancing measures, business closures) exerted substantial effects both on substance use patterns and treatment seeking for substance use disorders (SUD) (3). These disruptions created complex, potentially reinforcing cycles, as SUD may perpetuate NEET status (3). Employment upheavals also varied by industry which led to significant disparities among vulnerable socio-economic populations who were disproportionately impacted by pandemic-related job losses and substance-related harms (4, 5). Therefore, there is an urgent public health need to better understand the demographic profile, substance use behaviors, and SUD treatment utilization patterns of NEET men in the wake of the COVID-19 pandemic (6). These data can help policymakers and public health officials develop targeted interventions to address the complex relationship between NEET status and SUD while supporting workforce reintegration.

Multiple studies have identified bi-directional links between substance use disorders and NEET status among men. In a national longitudinal cohort, Manhica et al., demonstrated that NEET status among young adults (ages 17-24) was associated with an increased risk of later SUD diagnosis among males 25 and older (7, 8). In a study of 4,758 young Swiss men in their early 20s, previous cannabis use, and daily tobacco smoking predicted an increased likelihood of being NEET over 15 months (9). Rodwell demonstrated that adolescents (14–15) who used cannabis frequently (defined as at least weekly use) had a 74% higher likelihood of being NEET in young adulthood (24–25) compared to those who used cannabis infrequently or not at all. Although Rodwell’s study included both males and females, males had higher rates of frequent cannabis use during adolescence (19.3% of males versus 11.9% of females). Finally, Compton et al., showed that unemployed men and women aged 18 and older (defined as without a job in the past four weeks) had 40% higher rates of heavy alcohol use and 79% higher rates of illicit drug use than employed individuals (10). However, existing research on NEET men has typically focused on young adults and frequently lacked diverse samples, limiting generalizability to the context of the COVID-19 pandemic. Although the link between substance use and NEET status in men is robust, specific knowledge about substance use patterns among NEET men during this period remains limited.

Pandemic-related social and economic disruptions reshaped both substance-use behaviors and pathways to treatment which may have lasting implications (3). The US experienced an average increase in alcohol use (11), drug overdose rates and SUD hospitalizations (12, 13), and a decrease in tobacco use (14), with mixed evidence of increases in cannabis use (15). The COVID-19 pandemic created barriers to SUD treatment seeking through social distancing measures, but the relaxation of regulations on telehealth and medications for opioid use disorder may have helped counter forces that reduced treatment access (16, 17). Evidence on changes in health services access for SUD are varied and fragmented across different study settings (18). Moreover, these changes may have had disproportionate impacts among vulnerable populations such as NEET men. Therefore, understanding their specific behavioral patterns during this period offers critical insights into responses to large-scale societal stressors. For example, changes in the type of substances used may signal increased need for workforce training and program development for specific treatment strategies (e.g. medications for opioid use disorder, contingency management). Therefore, characterizing substance use and treatment utilization patterns using national data from this timeframe is essential for developing targeted and effective interventions to enhance public health and enhance workforce integration.

To help fill these knowledge gaps, we examined sociodemographic factors, substance use patterns, and SUD treatment utilization among working-age male participants in the 2022 National Survey on Drug Use and Health (NSDUH). Our primary objectives are to: (1) characterize substance-specific use patterns among working-age NEET vs non-NEET men, (2) examine their patterns of SUD treatment utilization, and (3) identify specific demographic variables for potential targeted interventions. These data can help develop policies and programs designed to help NEET men re-enter the workforce or education, mitigate the negative SUD-related consequences of NEET status, and increase national preparedness for future labor shocks (19–21).

## 2. Methods

### 2.1 Data source

We analyzed data from the 2022 National Survey on Drug Use and Health (NSDUH), an annual cross-sectional federal government survey of non-institutionalized individuals in the United States. The NSDUH obtains a nationally representative probability sample through a complex survey design for all 50 states and the District of Columbia (22). Surveys are administered via computer-assisted interviewing and audio computer-assisted self-interviewing, though self-report data may be subject to recall and social desirability bias, particularly for sensitive substance use behaviors. The analytic sample included 27,047 men of working age (18-64 years) from the 2022 NSDUH dataset. The NSDUH’s weighted interview response rates are between 60.4% and 69.3%. To address potential nonresponse bias, the survey implements demographic-based weight adjustments, comprehensive nonresponse bias analyses, and targeted follow-up with nonrespondents (22, 23). The study used publicly available and de-identified data and was exempt from IRB review.

### 2.2 Measures

The primary outcome was NEET status, defined as : 1) not working in the past year, 2) not currently enrolled in school or training programs, and 3) not retired. This definition captures longer-term disengagement from work and education rather than short-term unemployment. NEET status was coded as a binary variable. Cases with missing data on employment, education, or retirement status were excluded from analyses. All other variables had complete data due to NSDUH’s imputation procedures. We considered the following demographic characteristics: age (18-25, 26-34, 35-49, and 50-64 years), race/ethnicity (Non-Hispanic (NH) Asian, NH Black/African-American, Hispanic, NH Native American/Alaskan Native, NH Native Hawaiian/Pacific Islander, NH Multiracial, and NH White), education (less than high school, high school graduate, some college, and college graduate), annual family income (<$20,000, $20,000-$49,999, $50,000-$74,999, and >$75,000), relationship status (never married, married, and unmarried with the latter including widowed, divorced and separated individuals), and urbanicity (large metropolitan, small metropolitan, and non-metropolitan).

Past-month substance use was assessed for each substance through self-report. The specific substances included alcohol, ecstasy/molly, LSD, cannabis, cocaine, heroin, fentanyl, methamphetamine, prescription opioids, prescription stimulants, prescription pain relievers and tranquilizers/sedatives. Misuse was defined as use of prescription drugs in any way a doctor did not direct (23). We assessed past-month nicotine dependence defined by NSDUH using the Nicotine Dependence Syndrome Scale (24). We further assessed substance use frequencies of cannabis, heroin, methamphetamine and alcohol. SUD outcomes (i.e., past year disorder) for alcohol and drugs were based on criteria from the DSM-5 (25). Substance use treatment was categorized as a binary variable that included receiving treatment for alcohol or drug use at any location, such as a hospital, rehabilitation facility, mental health center, emergency room, or doctor’s office, including phone or video treatment.

### 2.3 Statistical analysis

We estimated the prevalence of NEET status among working-age men and compared demographic characteristics, substance use behaviors, and SUD treatment utilization patterns between NEET and non-NEET men. Bivariable comparisons were conducted using Rao-Scott chi-square to determine differences for each of these conditions with respect to NEET status. To examine the association between NEET status and our outcomes of interest, we developed logistic regression models and adjusted for selected demographic characteristics (age, race/ethnicity, marital status, and urbanicity).

To account for potential family-wise error resulting from multiple comparisons, we used familywise Bonferroni statistical correction for the 7 demographic variables (α = 0.007; 0.05/7) and the 11 outcomes variables (α = 0.0045; 0.05/11) to determine significance. We also ensured that multicollinearity was not present. Weights were included in all analyses and R studio version 4.3.2 was used to analyze data (26). The analysis incorporated NSDUH’s complex survey design using cluster weights, strata weights, and calibrated person-level weights (27).

## 3. Results

### 3.1 Sample characteristics

The largest proportions of respondents were heterosexual (88.9%, 95% CI: 87.9-89.8), non-Hispanic White (58.6%, 95% CI: 56.7-61.0), aged 35-49 (31.5%, 95% CI: 30.5-32.5) or 50-64 (30.6%, 95% CI: 29.1-32.2), married, (45.1% CI: 43.6-46.6), and resided in large metropolitan areas (55.6%, 95% CI: 53.3-57.9). An estimated 11.1% (95% CI: 10.1-12.1) of working aged (18–64) males in the US were not in employment, education or training (i.e., NEET) representing an estimated 10.6 million individuals (95% CI: 9.7-11.6 million). Table 1 presents demographic characteristics according to NEET status. Significant racial/ethnic disparities were observed with Non-Hispanic Black men showing higher NEET rates (18.5% vs 11.5%) and Non-Hispanic Asian men showing lower rates (1.8% vs 6.9%). Individuals reporting gay sexual orientation had significantly higher NEET rates compared to individuals reporting heterosexual orientation (7.5% vs 3.7%). Compared to non-NEET men, NEET men were significantly more likely to be older (46.6% aged 50-64 vs 28.7%), have lower income (45.9% earning <$20,000 vs 10.4%), and less likely to be married (29.5% vs 47.3%). They were also less likely to be college graduates (11.0% vs 34.0%) and more likely to live in non-metropolitan areas (17.6% vs 11.4%). All demographic differences were statistically significant (ps<0.007).

**Table 1.**
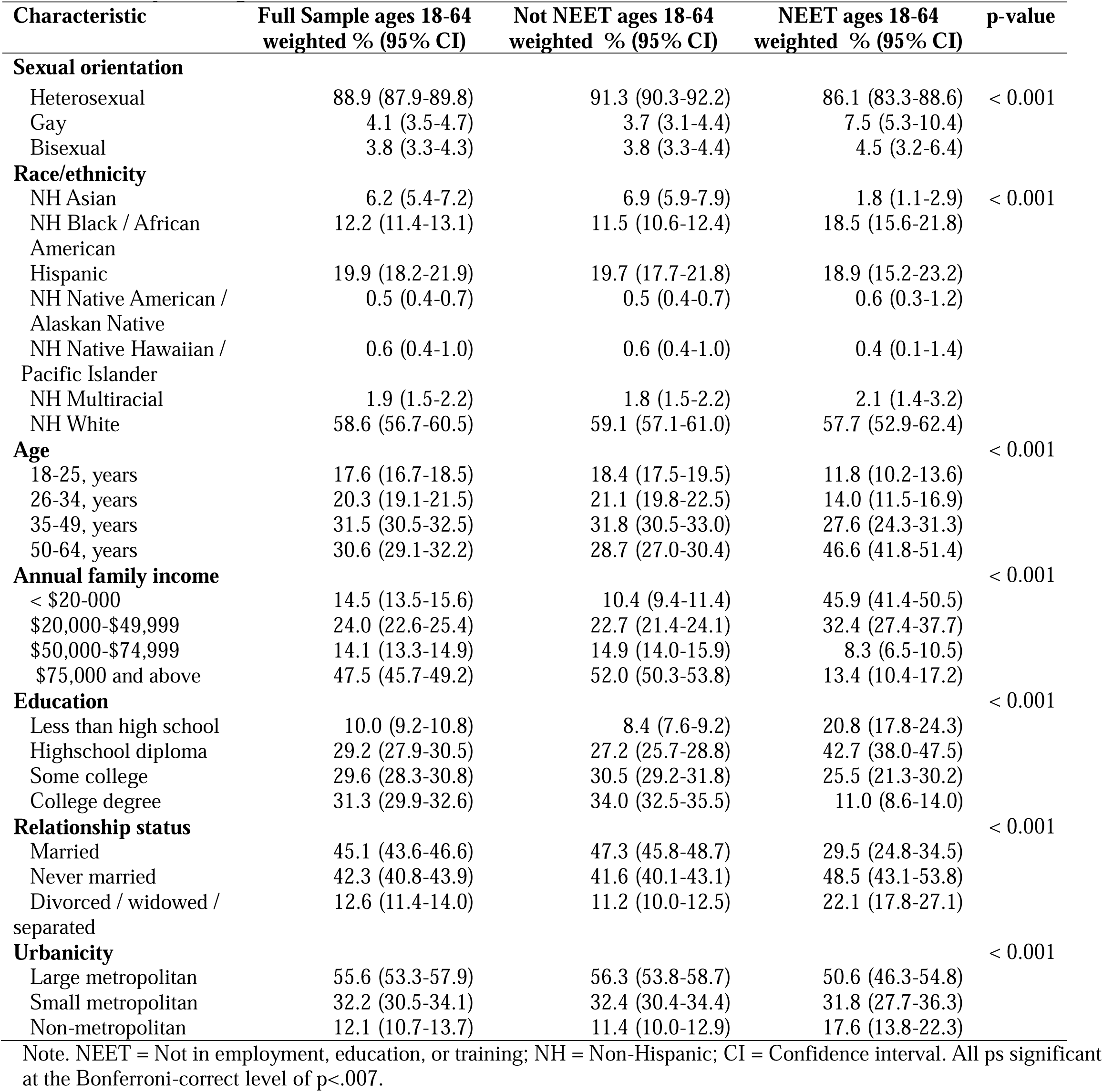
Demographic Characteristics of Working-Aged Men (18-64y) by NEET Status, United States, 2022.

### 3.2 Primary analyses

As shown in Table 2, NEET men had a distinctly different substance use and treatment profile than their non NEET counterparts. NEET men were more than twice as likely to have received any past-year substance use treatment (4.6 % vs 1.8 %; p = 0.008), and had higher rates of several substance use disorders including methamphetamine (3.5 % vs 0.8 %; p < 0.001), prescription opioid (5.5 % vs 1.7 %; p < 0.001) and tranquilizer/sedative use disorders (2.4 % vs 0.7 %; p < 0.001). NEET men were three times more likely to report opioid use disorder (heroin or prescription opioid) than non-NEET men (6.2 % vs 2.0 %; p < 0.001). Surprisingly, NEET men had significantly lower rates of alcohol use disorder compared to non-NEET men (11.9 % vs 16.3 %; p = 0.002). The overall prevalence of any past year SUD did not differ significantly between groups (27.2 % vs 24.5 %; p = 0.117).

**Table 2.**
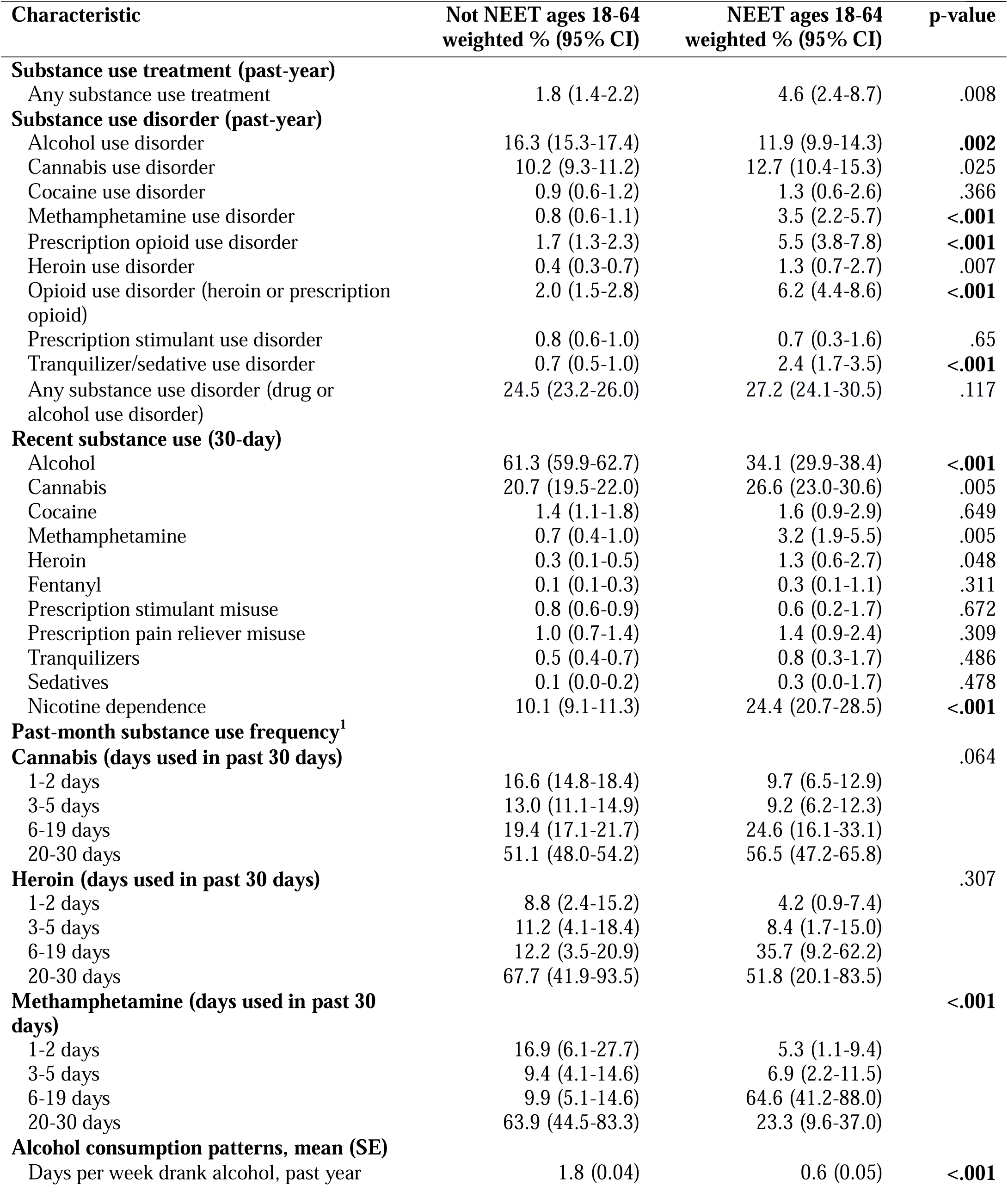
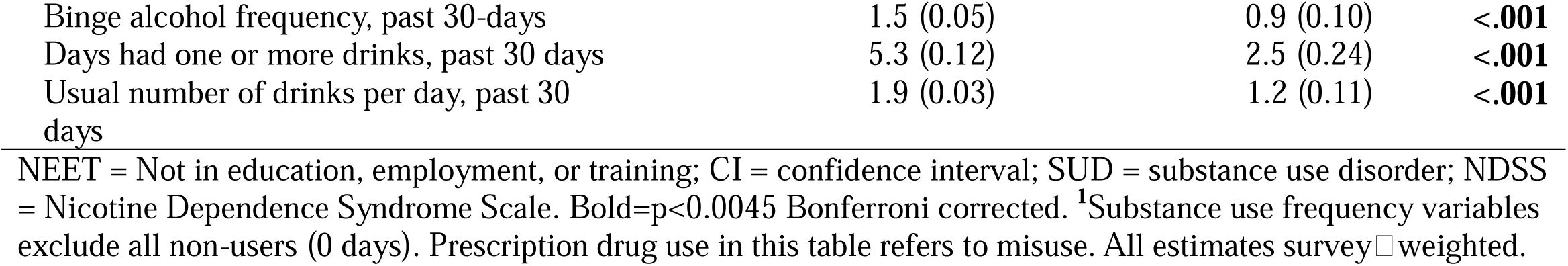
Substance Use Treatment, Disorders, and Recent Use Among Working □ Aged Men (18□64□y) by NEET Status, United States, 2022 National Survey on Drug Use and Health.

When looking at recent use, NEET men showed significant differences in nicotine dependence and measures of alcohol consumption but fewer differences regarding other substances. Specifically, NEET demonstrated more than twice the rate of nicotine dependence in the past 30 days compared to non-NEET men (10.1% vs. 24.4%, p<0.001), but reported fewer binge drinking episodes (0.89 vs. 1.48 days, p<0.001), fewer days drinking per week (0.61 vs. 1.81 days, p<0.001), and fewer drinks per day (1.20 vs. 1.91, p<0.001). Past 30-day alcohol consumption was significantly lower in NEET men (61.3% vs 34.1%, p < 0.001). Frequency distributions suggest that when NEET men used methamphetamine, they are less likely to be either casual users (1-2 days: 5.3% vs 16.9%) or daily users (20-30 days: 23.3% vs 63.9%) and show higher use rates at intermediate frequencies (6-19 days: 64.6% vs 9.9%; p<0.001).“

In our adjusted models (Table 3), with all else being equal, working-aged NEET men were at lower odds of past-year alcohol use disorder but substantially increased odds of several SUDs. After applying the Bonferroni corrected α = 0.0045, NEET status was associated with lower odds of alcohol use disorder (aOR = 0.68, 95 % CI = 0.54-0.85), and higher odds of cannabis use disorder (aOR = 1.40, 95 % CI = 1.13-1.74), methamphetamine use disorder (aOR = 3.10, 95 % CI = 1.82-5.28), prescription pain reliever use disorder (aOR = 2.88, 95 % CI = 1.82-4.53), any opioid use disorder (heroin or prescription; aOR = 2.68, 95 % CI = 1.71-4.20, and prescription tranquilizer/sedative use disorder (aOR = 3.54, 95 % CI = 1.97-6.37). No significant differences emerged for cocaine, heroin, prescription stimulant, or any drug and alcohol use disorder, and the association with past year substance use treatment engagement fell below the Bonferroni-adjusted threshold for significance. Overall, the adjusted findings highlight a distinct substance use pattern in which NEET men carry disproportionate risks for high harm prescription and illicit drug disorders while exhibiting comparatively lower alcohol involvement.

**Table 3.**
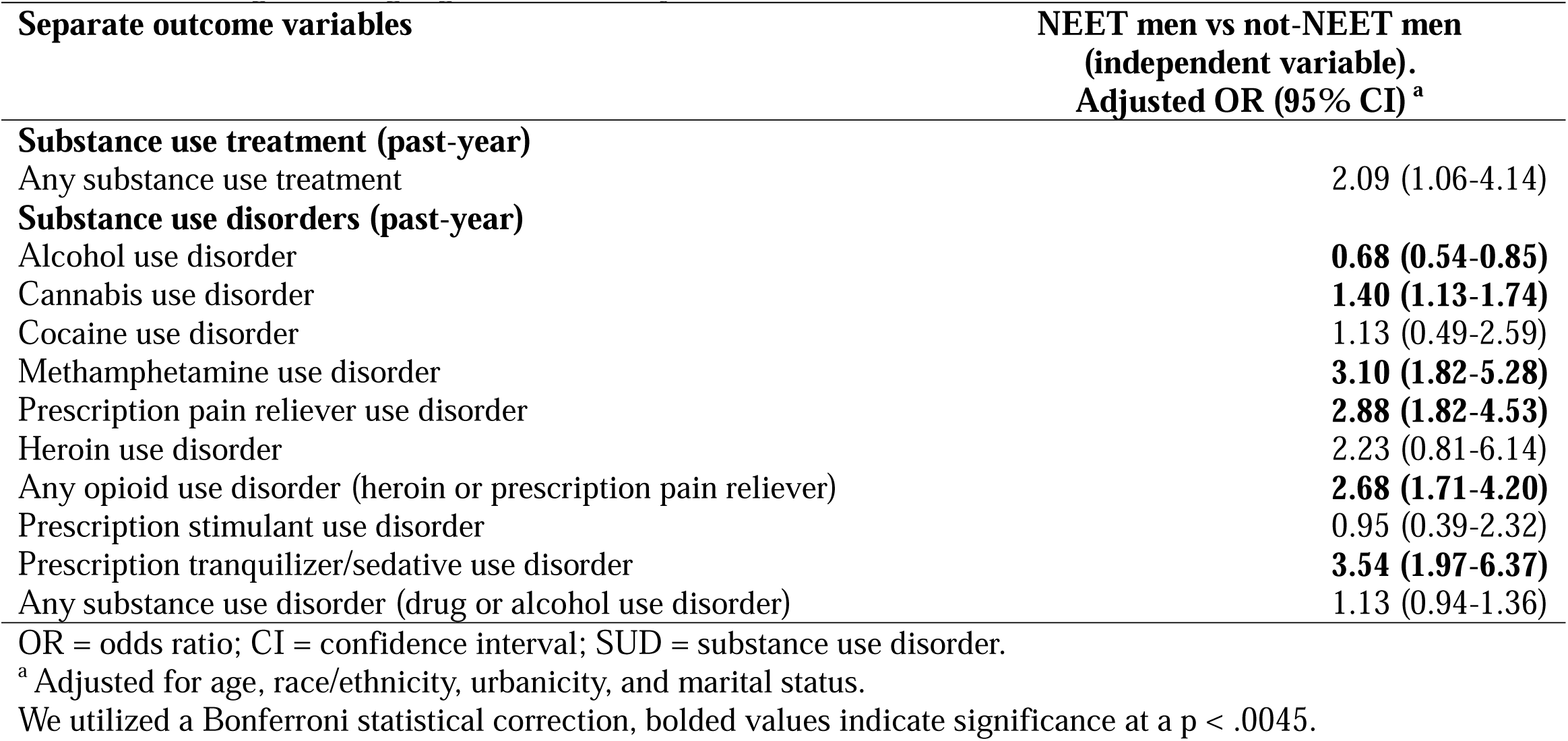
Multivariable Models Examining NEET Status as a Risk Factor for Past-Year Substance Use Disorders Among Working-Aged Men (18-64y) in the United States, 2022.

## 4. Discussion

### 4.1 Principal findings

In this nationally representative study, NEET men had substantially higher odds of prescription tranquilizer/sedative use disorder, methamphetamine use disorder, and opioid use disorder compared to their counterparts. We estimated that 11.1% of working-aged men (18–64) were not engaged in employment, education or training in 2022 during the COVID-19 pandemic. This is remarkable given that the U.S. enjoyed historically low rates of unemployment (less than 4%) at the time of data collection (28). Our study adds to the literature by providing a more comprehensive picture of the NEET population that shows an overrepresentation of middle-aged and older adults (29). Consistent with past research, we found a high prevalence of past-year SUDs among NEET men, but past 30-day use of illicit substances was similar among both groups. Importantly, NEET men showed strikingly lower rates of alcohol use and higher rates of nicotine dependence. Despite NEET men being disproportionately in rural areas where SUD treatment services tend to in short supply, they had slightly higher prevalence of SUD treatment engagement. These findings indicate that public health interventions should be tailored by substance type (e.g., prescription drugs, methamphetamine) and designed with age- and culturally-appropriate components for this vulnerable population.

Our estimate that 11.1% of men (18–64) were NEET in 2022 closely aligns with previous research indicating that 11% of prime-aged men (aged 25-54) were not participating in the labor force in 2019 (Bureau of Labor Statistics, 2023; Eberstadt, 2022). NEET men also had significantly higher past-year prevalence of several SUDs which is consistent with previous work that has established independent associations between NEET status and substance use (7, 19, 30).

In contrast with previous literature, NEET men had significantly lower alcohol use among several key measures including fewer binge drinking episodes, fewer drinking days per week, fewer days with any alcohol consumption in the past month, and fewer drinks per day. This finding contrasts with an earlier study using the same NSDUH dataset which found significantly higher rates of heavy alcohol use and alcohol use disorders among unemployed individuals between 2002 and 2010 (10). However, Compton et al. defined unemployed as those without a job in the last four weeks whereas our study looked at longer-term unemployment which was defined as at 52 weeks. Although alcohol consumption frequency increased in the general population during the COVID-19 pandemic (11, 31), this trend varied among different demographic groups (32, 33).

Our finding of lower alcohol use among NEET men aligns with a “polarization effect” in alcohol consumption patterns during the COVID-19 pandemic. A study of Swiss men demonstrated that those reporting higher COVID-19-related health concerns and/or financial anxieties reported a decrease in their alcohol consumption, while those with fewer such concerns reported an increase (34). A large study of over 30,000 adults in the UK found a similar polarization based on socioeconomic position; individuals with low income were more likely to reduce their drinking whereas those with high income were more likely to drink more during the pandemic (35). Given that nearly half of the NEET men in our study earned less than $20,000 annually and were over the age of 50, it is likely they experienced the heightened health and financial vulnerabilities that led to a reduction in alcohol consumption as a protective or cost-saving response to pandemic-related stressors.

NEET men had substantially higher rates of past-year SUDs for multiple substances, but measures of past 30-day substance use were not significantly different between the two groups. This might suggest that some individuals reduced or ceased their substance use after becoming NEET due to financial constraints. This discrepancy might also indicate that comparable use of illicit substances might lead to more severe levels of functional impairment in the context of long-term unemployment, socioeconomic differences and the COVID-19 pandemic (36). Social distancing measures, telehealth prescribing of prescription medications, lower prices for synthetic drugs and mental health co-morbidity may partially explain this specific substance use profile (13, 37–39). For example, NEET status was shown to be associated with severe symptoms of anxiety and moderate/severe symptoms of depression and concurrent mental health and substance problems (30, 40). Our population’s older age and over-representation of Non-Hispanic Black males may also partially explain our findings due to increased intersectional vulnerability (41, 42).

In our sample, NEET status was disproportionately concentrated among older working age adults, which contrasts with much of the existing literature that predominantly focuses on adolescent and young adult NEET populations (Mawn et al., 2017; O’Dea et al., 2014; Rodwell et al., 2018; Stea et al., 2024). This suggests that COVID-19 may have uniquely affected older adult workers in terms of employment challenges compared to previous economic downturns (Li and Mutchler, 2021). It could also be that older men have been using substances for more years, which may have increased the severity of their disorder and simultaneously entrenched their NEET status (Kuerbis, 2020; Silver et al., 2022). The significant overrepresentation of men who identify as Non-Hispanic Black in the NEET population underscores the need for culturally tailored interventions. NEET men were significantly more likely to lack a high school diploma and far less likely to hold college degrees. These demographic patterns suggest disparities around the impact of COVID-19 on employment sectors especially among those requiring in-person work and those employing men without college credentials (Moen et al., 2020). The concentration of NEET status among older working age adult men represents a significant shift from the traditional view of NEET as a youth-centered problem.

Although NEET men showed higher rates of treatment seeking, this difference attenuated after adjustment which suggests that interventions should prioritize enhancing treatment effectiveness and retention as well as substance-specific programs. Previous studies show mixed results, with some indicating a treatment gap among long-term unemployed individuals, while others suggest unemployment rates significantly correlate with substance use treatment admissions (43, 44). Potential mechanisms for the attenuation include differential treatment access by rurality, demographic profile (older vs younger), and the availability of substance specific treatment options (e.g. methamphetamines).

The high prevalence of illicit substance use among the NEET population is cause for concern due to the potential for serious harms related to hospitalizations and fatal overdose especially among vulnerable populations (45). For example, hospitalizations for stimulant use increased by 42.6 % for adults aged 55-64 in California from 2017-2021 (46). Beginning in 2020, over 50% of all drug overdose deaths among adults aged 25-64 are estimated to have occurred among males with at most a high school-level education (47). During the COVID-19 pandemic, Black individuals experienced the largest percentage increase in drug overdose mortality (48.8%), surpassing overdose rates among White individuals for the first time since 1999 (48). These data suggest that a high proportion of NEET men in our sample are at increased risk of mortality due to their demographic and substance use profiles and likely account for a disproportionate number of overdose fatalities.

### 4.2 Implications

NEET men during the COVID-19 pandemic display unique substance use behaviors that have higher risk profiles which warrant immediate attention. Our findings highlight the urgent need for an integrated model of treatment for NEET men that addresses substance use, employment, and socio-economic barriers to workforce reentry and risk mitigation. For example, increasing educational attainment and improving economic conditions could have the potential to decrease unemployment rates and reduce substance use disorders simultaneously. Promising approaches include substance-specific programs targeting prevalent drugs among NEET men (e.g. methamphetamine, opioids), contingency management interventions that simultaneously address employment and substance use, re-education programs that target low-income communities or justice-involved individuals, and policy changes that extend health coverage for individuals transitioning out of employment to maintain treatment continuity (49). Focusing on enhancing work-related recovery capital for NEET men engaged in treatment may have indirect benefits for SUD outcomes (50); professional treatment programs (e.g. physician’s health programs) have high rates of success (51).

Evidence-based interventions such as overdose rescue medications and medications for opioid use disorder are critical tools for treatment and should be and made easily available for low-income and uninsured populations and accessible in a wide variety of settings (52, 53). The widespread use of synthetic drugs in the post-COVID era, and the new epidemic of stimulant and opioid co-use make these interventions critically important (54). Early prevention strategies aimed at reducing harmful substance use among unemployed men is warranted, especially when considering that stimulant use disorders have no FDA-approved medication-based treatment options. Future studies should consider adapting substance use prevention programs tailored to unemployed youth to middle-aged and older adults of different cultural backgrounds (55). Because the COVID-19 pandemic likely influenced our findings related to the NEET population, follow-up studies are warranted.

### 4.3 Limitations

Our study is limited due to the self-report nature of the survey which is therefore susceptible to recall and social desirability bias. Additionally, self-report of drug class used may lead to misidentification of specific substances. Multiple testing could be a limitation, but test results (ps < 0.0045) remained significant in light of a Bonferroni correction (alpha = 0.0045). In addition, our study is limited to community-living NEET men aged 18-64 and may not be representative of certain vulnerable subpopulations such as individuals experiencing homelessness who do not use shelters or those in institutional settings. Also, the NSDUH does not include a diagnostic interview; responses to survey questions are not a direct clinical diagnosis. However, proxy diagnoses in the NSDUH are used extensively in the literature and have been validated in previous studies. Additionally, while our study examines potential COVID-19 impacts on NEET men’s substance use patterns, the cross-sectional nature of our data limits causal inference regarding pandemic-related effects.

## 5. Conclusion

More than one in ten U.S. working-aged men were NEET in 2022, and they showed elevated risk of tranquilizer/sedative, methamphetamine, and prescription-opioid and use disorders while showing unexpectedly lower alcohol use. These findings call for multi-level evidence-based interventions that integrate substance-specific treatments (e.g., MOUD, contingency management for stimulants) with vocational assistance, and healthcare policies that maintain insurance coverage during periods of long-term unemployment; targeted interventions should be age-appropriate and culturally tailored.

## Data Availability

All data produced are available online at: https://www.samhsa.gov/data/data-we-collect/nsduh-national-survey-drug-use-and-health/datafiles

## Declarations

## Availability of Data and Materials

The data used in this study came from publicly available datasets from the National Survey on Drug Use and Health (NSDUH). These datasets can be accessed through the Substance Abuse and Mental Health Data Archive (SAMHDA) at https://datafiles.samhsa.gov/ after a brief application process. Additional data from these analyses are available from the authors upon reasonable request

## Authors’ Contributions

The individual contributions of each author are specified using the CRediT (Contributor Roles Taxonomy) author statement.

**Keith Humphreys:** Conceptualization, Investigation, Writing - Original Draft, Writing - Review & Editing, Supervision, and Funding acquisition.

**Wayne Kepner:** Conceptualization, Methodology, Formal analysis, Investigation, Resources, Data Curation, Writing - Original Draft, Writing - Review & Editing, Project administration, and Funding acquisition.

## Ethics Approval and Consent to Participate

Not applicable.

## Consent for Publication

Not applicable.

## Acknowledgements

Not applicable.

