## Supplementary material for "Substance use and treatment utilization patterns of working-age American men who were not in employment, education, or training (NEET) during the COVID-19 pandemic": Table 1

| Table 1. Demographic Characteristics of Working-Aged Men (18-64y) by NEET Status, United States, 2022 National Survey on Drug Use and Health | | | | |
| --- | --- | --- | --- | --- |
| Characteristic | **Full Sample ages 18-64 weighted % (95% CI)** | **Not NEET ages 18-64 weighted % (95% CI)** | **NEET ages 18-64 weighted % (95% CI)** | **P value** |
| Sexual orientation |  |  |  |  |
| Heterosexual | 88.9 (87.9-89.8) | 91.3 (90.3-92.2) | 86.1 (83.3-88.6) | < 0.001 |
| Gay | 4.1 (3.5-4.7) | 3.7 (3.1-4.4) | 7.5 (5.3-10.4) |  |
| Bisexual | 3.8 (3.3-4.3) | 3.8 (3.3-4.4) | 4.5 (3.2-6.4) |  |
| Race/ethnicity |  |  |  |  |
| NH Asian | 6.2 (5.4-7.2) | 6.9 (5.9-7.9) | 1.8 (1.1-2.9) | < 0.001 |
| NH Black / African American | 12.2 (11.4-13.1) | 11.5 (10.6-12.4) | 18.5 (15.6-21.8) |  |
| Hispanic | 19.9 (18.2-21.9) | 19.7 (17.7-21.8) | 18.9 (15.2-23.2) |  |
| NH Native American / Alaskan Native | 0.5 (0.4-0.7) | 0.5 (0.4-0.7) | 0.6 (0.3-1.2) |  |
| NH Native Hawaiian / Pacific Islander | 0.6 (0.4-1.0) | 0.6 (0.4-1.0) | 0.4 (0.1-1.4) |  |
| NH Multiracial | 1.9 (1.5-2.2) | 1.8 (1.5-2.2) | 2.1 (1.4-3.2) |  |
| NH White | 58.6 (56.7-60.5) | 59.1 (57.1-61.0) | 57.7 (52.9-62.4) |  |
| Age |  |  |  | < 0.001 |
| 18-25, years | 17.6 (16.7-18.5) | 18.4 (17.5-19.5) | 11.8 (10.2-13.6) |  |
| 26-34, years | 20.3 (19.1-21.5) | 21.1 (19.8-22.5) | 14.0 (11.5-16.9) |  |
| 35-49, years | 31.5 (30.5-32.5) | 31.8 (30.5-33.0) | 27.6 (24.3-31.3) |  |
| 50-64, years | 30.6 (29.1-32.2) | 28.7 (27.0-30.4) | 46.6 (41.8-51.4) |  |
| Annual family income |  |  |  | < 0.001 |
| < $20-000 | 14.5 (13.5-15.6) | 10.4 (9.4-11.4) | 45.9 (41.4-50.5) |  |
| $20,000-$49,999 | 24.0 (22.6-25.4) | 22.7 (21.4-24.1) | 32.4 (27.4-37.7) |  |
| $50,000-$74,999 | 14.1 (13.3-14.9) | 14.9 (14.0-15.9) | 8.3 (6.5-10.5) |  |
| $75,000 and above | 47.5 (45.7-49.2) | 52.0 (50.3-53.8) | 13.4 (10.4-17.2) |  |
| Education |  |  |  | < 0.001 |
| Less than high school | 10.0 (9.2-10.8) | 8.4 (7.6-9.2) | 20.8 (17.8-24.3) |  |
| Highschool diploma | 29.2 (27.9-30.5) | 27.2 (25.7-28.8) | 42.7 (38.0-47.5) |  |
| Some college | 29.6 (28.3-30.8) | 30.5 (29.2-31.8) | 25.5 (21.3-30.2) |  |
| College degree | 31.3 (29.9-32.6) | 34.0 (32.5-35.5) | 11.0 (8.6-14.0) |  |
| Relationship status |  |  |  | < 0.001 |
| Married | 45.1 (43.6-46.6) | 47.3 (45.8-48.7) | 29.5 (24.8-34.5) |  |
| Never married | 42.3 (40.8-43.9) | 41.6 (40.1-43.1) | 48.5 (43.1-53.8) |  |
| Divorced / widowed / separated | 12.6 (11.4-14.0) | 11.2 (10.0-12.5) | 22.1 (17.8-27.1) |  |
| Urbanicity |  |  |  | < 0.001 |
| Large metropolitan | 55.6 (53.3-57.9) | 56.3 (53.8-58.7) | 50.6 (46.3-54.8) |  |
| Small metropolitan | 32.2 (30.5-34.1) | 32.4 (30.4-34.4) | 31.8 (27.7-36.3) |  |
| Non-metropolitan | 12.1 (10.7-13.7) | 11.4 (10.0-12.9) | 17.6 (13.8-22.3) |  |

Note. NEET = Not in employment, education, or training; NH = Non-Hispanic; CI = Confidence interval. All ps significant at the Bonferroni-correct level of p<.007.
