## Supplementary material for "Substance use and treatment utilization patterns of working-age American men who were not in employment, education, or training (NEET) during the COVID-19 pandemic": Table 2

| Table 2. Substance Use Treatment, Disorders, and Recent Use Among Working‑Aged Men (18‑64 y) by NEET Status, United States, 2022 National Survey on Drug Use and Health | | | |
| --- | --- | --- | --- |
| Characteristic | **Not NEET ages 18-64 weighted % (95% CI)** | **NEET ages 18-64 weighted % (95% CI)** | **p-value** |
| Substance use treatment (past-year) |  |  |  |
| Any substance use treatment | 1.8 (1.4-2.2) | 4.6 (2.4-8.7) | .008 |
| Substance use disorder (past-year) |  |  |  |
| Alcohol use disorder | 16.3 (15.3-17.4) | 11.9 (9.9-14.3) | **.002** |
| Cannabis use disorder | 10.2 (9.3-11.2) | 12.7 (10.4-15.3) | .025 |
| Cocaine use disorder | 0.9 (0.6-1.2) | 1.3 (0.6-2.6) | .366 |
| Methamphetamine use disorder | 0.8 (0.6-1.1) | 3.5 (2.2-5.7) | **<.001** |
| Prescription opioid use disorder | 1.7 (1.3-2.3) | 5.5 (3.8-7.8) | **<.001** |
| Heroin use disorder | 0.4 (0.3-0.7) | 1.3 (0.7-2.7) | .007 |
| Opioid use disorder (heroin or prescription opioid) | 2.0 (1.5-2.8) | 6.2 (4.4-8.6) | **<.001** |
| Prescription stimulant use disorder | 0.8 (0.6-1.0) | 0.7 (0.3-1.6) | .65 |
| Tranquilizer/sedative use disorder | 0.7 (0.5-1.0) | 2.4 (1.7-3.5) | **<.001** |
| Any substance use disorder (drug or alcohol use disorder) | 24.5 (23.2-26.0) | 27.2 (24.1-30.5) | .117 |
| Recent substance use (30-day) |  |  |  |
| Alcohol | 61.3 (59.9-62.7) | 34.1 (29.9-38.4) | **<.001** |
| Cannabis | 20.7 (19.5-22.0) | 26.6 (23.0-30.6) | .005 |
| Cocaine | 1.4 (1.1-1.8) | 1.6 (0.9-2.9) | .649 |
| Methamphetamine | 0.7 (0.4-1.0) | 3.2 (1.9-5.5) | .005 |
| Heroin | 0.3 (0.1-0.5) | 1.3 (0.6-2.7) | .048 |
| Fentanyl | 0.1 (0.1-0.3) | 0.3 (0.1-1.1) | .311 |
| Prescription stimulant misuse | 0.8 (0.6-0.9) | 0.6 (0.2-1.7) | .672 |
| Prescription pain reliever misuse | 1.0 (0.7-1.4) | 1.4 (0.9-2.4) | .309 |
| Tranquilizers | 0.5 (0.4-0.7) | 0.8 (0.3-1.7) | .486 |
| Sedatives | 0.1 (0.0-0.2) | 0.3 (0.0-1.7) | .478 |
| Nicotine dependence (NDSS) | 10.1 (9.1-11.3) | 24.4 (20.7-28.5) | **<.001** |
| Past-month substance use frequency^1^ |  |  |  |
| Cannabis (days used in past 30 days) |  |  | .064 |
| 1-2 days | 16.6 (14.8-18.4) | 9.7 (6.5-12.9) |  |
| 3-5 days | 13.0 (11.1-14.9) | 9.2 (6.2-12.3) |  |
| 6-19 days | 19.4 (17.1-21.7) | 24.6 (16.1-33.1) |  |
| 20-30 days | 51.1 (48.0-54.2) | 56.5 (47.2-65.8) |  |
| Heroin (days used in past 30 days) |  |  | .307 |
| 1-2 days | 8.8 (2.4-15.2) | 4.2 (0.9-7.4) |  |
| 3-5 days | 11.2 (4.1-18.4) | 8.4 (1.7-15.0) |  |
| 6-19 days | 12.2 (3.5-20.9) | 35.7 (9.2-62.2) |  |
| 20-30 days | 67.7 (41.9-93.5) | 51.8 (20.1-83.5) |  |
| Methamphetamine (days used in past 30 days) |  |  | **<.001** |
| 1-2 days | 16.9 (6.1-27.7) | 5.3 (1.1-9.4) |  |
| 3-5 days | 9.4 (4.1-14.6) | 6.9 (2.2-11.5) |  |
| 6-19 days | 9.9 (5.1-14.6) | 64.6 (41.2-88.0) |  |
| 20-30 days | 63.9 (44.5-83.3) | 23.3 (9.6-37.0) |  |
| Alcohol consumption patterns, mean (SE) |  |  |  |
| Days per week drank alcohol, past year | 1.8 (0.04) | 0.6 (0.05) | **<.001** |
| Binge alcohol frequency, past 30-days | 1.5 (0.05) | 0.9 (0.10) | **<.001** |
| Days had one or more drinks, past 30 days | 5.3 (0.12) | 2.5 (0.24) | **<.001** |
| Usual number of drinks per day, past 30 days | 1.9 (0.03) | 1.2 (0.11) | **<.001** |
| NEET = Not in education, employment, or training; CI = confidence interval; SUD = substance use disorder; NDSS = Nicotine Dependence Syndrome Scale. Bold=p<0.0045 Bonferroni corrected. ^1^Substance use frequency variables exclude all non-users (0 days). Prescription drug use in this table refers to misuse. All estimates survey‑weighted. | | | |
