## Supplementary material for "Substance use and treatment utilization patterns of working-age American men who were not in employment, education, or training (NEET) during the COVID-19 pandemic": Table 3

| Table 3. Multivariable Models Examining NEET Status as a Risk Factor for Past-Year Substance Use Disorders Among Working-Aged Men (18-64y) in the United States, 2022 | |
| --- | --- |
| Separate outcome variables | **NEET men vs not-NEET men (independent variable).  Adjusted OR (95% CI) ^a^** |
| Substance use treatment (past-year) |  |
| Any substance use treatment | 2.09 (1.06–4.14) |
| Substance use disorders (past-year) |  |
| Alcohol use disorder | **0.68 (0.54–0.85)** |
| Cannabis use disorder | **1.40 (1.13–1.74)** |
| Cocaine use disorder | 1.13 (0.49–2.59) |
| Methamphetamine use disorder | **3.10 (1.82–5.28)** |
| Prescription pain reliever use disorder | **2.88 (1.82–4.53)** |
| Heroin use disorder | 2.23 (0.81–6.14) |
| Any opioid use disorder (heroin or prescription pain reliever) | **2.68 (1.71–4.20)** |
| Prescription stimulant use disorder | 0.95 (0.39–2.32) |
| Prescription tranquilizer/sedative use disorder | **3.54 (1.97–6.37)** |
| Any substance use disorder (drug or alcohol use disorder) | 1.13 (0.94–1.36) |
| OR = odds ratio; CI = confidence interval; SUD = substance use disorder. ^a^ Adjusted for age, race/ethnicity, urbanicity, and marital status. We utilized a Bonferroni statistical correction, bolded values indicate significance at a p < .0045. | |
